# MAGNITUDE OF METABOLIC SYNDROME AND ITS ASSOCIATED FACTORS AMONG PATIENTS WITH TYPE 2 DIABETES MELLITUS WHO ARE ON FOLLOW-UP AT ASELLA REFERRAL AND TEACHING HOSPITAL DIABETIC CLINIC, ASELLA, OROMIA, ETHIPIA IN 2022

**DOI:** 10.64898/2026.11.18.26367698

**Authors:** Natnael Tegenu Mitiku, Leul Mekonnen Nigatu, Nebyu Yonas Shanka

**Author notes:** Corrsponding author.

## Abstract

**Background:** Type 2 Diabetes mellites and Metabolic syndrome are global public health problems, and their burden is rising, particularly in developing countries. Metabolic syndrome (syndrome X, insulin resistance syndrome) consists of a constellation of metabolic abnormalities that confer increased risk of cardiovascular disease (CVD) and diabetes mellitus. There are limited data on prevalence of metabolic syndrome in type II DM patients in Ethiopia and in the horn of Africa at large, and no research has been done in our institution on this topic.

**Objective:** To determine the prevalence of metabolic syndrome and its associated factors in patients with type 2 diabetes on follow up at Asella Referral and Teaching Hospital diabetes follow up clinic, Asella, Oromia, Ethiopia.

**Method:** Cross-sectional, prospective, hospital-based survey was conducted to assess the prevalence of metabolic syndrome and its associated factors in type II DM patients among those who visited diabetic follow-up clinic of Asella referral and teaching Hospital. All type II DM patients, who visited the diabetic follow-up clinic in the study period was included in the study. 285 participants were selected using systemic random sampling technique. Data was collected from patients and their chart and filled on a checklist by trained data collectors. Data was entered to epi-data cleared and analyzed by SPSS version 26.0 and Odds ratio with 95% CI and P-value of <0.05 was considered for declaring level of significance.

**Results:** Using the NCEP-ATP III criteria the magnitude of Metabolic syndrome was 75.1% in this study. Among the components of Metabolic syndrome high blood pressure was the commonest (70.9%) followed by low HDL Cholesterol (70.5%), high Triglyceride (46%) and high Waist circumference (12.6%) respectively. Female and Male had comparable prevalence of Metabolic syndrome and the study showed that 55.4% of participants were taking Lipid lowering medications. Multivariate logistic regression analysis showed four factors; frequency of taking Meat (AOR = 2.47; 95% CI = 1.323-4.6), Alcohol intake (AOR = 2.79; 95% CI = 1.146-6.806), total Cholesterol level (AOR = 2.72; 95% CI = 1.023-7.224) and LDL Cholesterol level (AOR = 2.31; 95% CI = (1.029-5.161) were significantly associated with Metabolic syndrome according to this study.

**Conclusion:** In this study the magnitude of Metabolic syndrome was found to be 75.1% and the associated factors were Alcohol intake, Frequency of taking Meat, Total and LDL Cholesterol level.

**Recommendation:** Counseling of Type 2 Diabetes Mellitus patients on the need of decreasing Alcohol and Meat consumption and physicians need to improve the habit of prescribing Lipid lowering medication.

## 1. Introduction

### 1.1 Background

Diabetes mellitus (DM) refers to a group of common metabolic disorders that share the phenotype of hyperglycemia. Several distinct types of DM are caused by a complex interaction of genetics and environmental factors. Depending on the etiology of the DM, factors contributing to hyperglycemia include reduced insulin secretion and disorders of Glucose utilization and production. There are two broad categories of DM, designated as either type 1 or type 2 DM. Type 2 DM is a heterogeneous group of disorders characterized by variable degrees of insulin resistance, impaired insulin secretion, and increased hepatic glucose production(1).

The metabolic syndrome (syndrome X, insulin resistance syndrome) consists of a constellation of metabolic abnormalities that confer increased risk of cardiovascular disease (CVD) and diabetes mellitus. Evolution of the criteria for the metabolic syndrome since the original definition by the World Health Organization in 1998 reflects growing clinical evidence and analysis by a variety of consensus conferences and professional organizations. The major features of the metabolic syndrome include central obesity, hypertriglyceridemia, low levels of high-density lipoprotein (HDL) cholesterol, hyperglycemia, and hypertension(1).

Nowadays, there are different institutions that set different diagnostic criteria for Metabolic Syndrome worldwide. These include the criteria by the World Health Organization (WHO), the National Cholesterol Education Program Adult Treatment Panel III (NCEP-ATP III), the harmonized criteria, and the International Diabetes Federation (IDF). The institutions have their own general agreement on the main component of the Metabolic Syndrome, but they differ based on the mandatory inclusion criteria.(2)

According to these expert groups, its main components are raised fasting plasma glucose, dyslipidemia and elevation of arterial blood pressure (BP), while abdominal obesity and/or insulin resistance (IR) have gained increasing attention as the core manifestations of the syndrome.(2) NCEP/ATP III requires at least three of the following:1. central obesity: waist circumference ≥ 102 cm (male), ≥ 88 cm (female), 2. Dyslipidemia: TG ≥ 1.7 mmol/L (150 mg/dl), 3. dyslipidemia: HDL-C < 40 mg/dL (male), < 50 mg/ dL (female)\, 4. blood pressure ≥ 130/85 mmHg, 5. fasting plasma glucose ≥ 6.1 mmol/l (110 mg/dl)(3).

The IDF consensus worldwide definition of the metabolic syndrome (2005) include, central obesity (defined as waist circumference with ethnicity specific values)and any two of the following: 1. Raised triglycerides: > 150 mg/dL (1.7 mmol/L), or specific treatment, 2. Reduced HDL cholesterol: < 40 mg/dL (1.03 mmol/L) in males, < 50 mg/dL (1.29 mmol/L) in females, or specific treatment for this lipid abnormality, 3. Raised blood pressure: systolic BP > 130 or diastolic BP >85 mm Hg, or under treatment, 4. Raised fasting plasma glucose: >100 mg/dL (5.6 mmol/L), or previously diagnosed type 2 diabetes. Waist circumferences with ethnicity specific values in IDF are as: 1. Male ≥94 cm and female ≥80 cm for European descent, 2. Male >90 cm and female >80 cm for South Asian (Chinese, Asian-Indian) descent, 3. Male >102 cm and female >88 cm for American descent(3).

### 1.2 Statement of the problem

The worldwide prevalence of DM has risen dramatically over the past two decades, from an estimated 30 million cases in 1985 to 415 million in 2017. Based on current trends, the IDF projects that 642 million individuals will have diabetes by the year 2040. Although the prevalence of both type 1 and type 2 DM is increasing worldwide, the prevalence of type 2 DM is rising much more rapidly, presumably because of increasing obesity, reduced activity levels as countries become more industrialized, and the aging of the population (1).

It is estimated that around 20-25% of the world’s adult population have Metabolic Syndrome and they are 3 times as likely to have a heart attack compared with people without the syndrome. The clustering CVD risk factors that illustrates Metabolic Syndrome is now considered to be the driving force for a new CVD epidemic and people with T2DM who have Metabolic Syndrome carry a much higher risk of CVD than those who have T2DM alone(4).

Diabetes mellitus (DM) and related complications are associated with long-term damage and failure of various organ systems. It induces changes in the microvasculature, causing extracellular matrix protein synthesis and capillary basement membrane thickening which are the pathognomonic features of diabetic microangiopathy, these changes in conjunction with advanced glycation end products oxidative stress, low grade inflammation and neovascularization of vasa vasorum can lead to vascular complications. Rapid urbanization and major changes in lifestyle could be driving this complications(5).

The issue of diabetes mellitus and metabolic syndrome is becoming a major concern in developing countries like Ethiopia, according to my hypothesis in Assela teaching and referral Hospital most of the outpatient cases and ward admitted cases are directly related or indirectly associated to metabolic syndrome or type II DM. This is related to urbanization which encompass change in dietary habit and limitation of activity or immobilization.

There are limited data on metabolic syndrome in Ethiopia and in the horn of Africa at large. No research has been done in our institution on this topic, Therefore, this study is helpful to the development of lifestyle modification, control and management of the metabolic syndrome. Finally, this will bring non-communicable disease in general and metabolic syndrome in particular into the clinical area, and research agenda.

### 1.3 Significance of the study

Applying the criteria for the metabolic syndrome serves as a simple and inexpensive tool for identifying patients at high risk for diabetes and coronary heart disease, particularly those who do not fall into traditional risk categories. Several independent physiological processes underlie the non-random risk-factor clustering that defines the metabolic syndrome, including insulin resistance, central obesity, dyslipidemia, impaired glucose tolerance, and hypertension. Other non-classic risk factors, such as abnormal oxidized low-density lipoprotein-cholesterol, adiponectin, and C-reactive protein levels, are highly correlated with the metabolic syndrome. Use of the metabolic syndrome criteria for assessment is comparable with other risk-scoring systems in accurately predicting cardiovascular disease risk and is simpler to implement in the clinic(6).

At Assela referral and teaching hospital diabetic clinic, I noticed that T II DM patients were not being screened adequately for metabolic syndrome. For example, Waist circumference is not being done at all and some basic investigations like lipid profile are lacking for majority of patients. For the above reason most of the patients are not getting important drugs like Statins and Aspirin, American diabetic association recommends use of statin for every diabetic patient age between 40-75 years with or without atherosclerotic cardiovascular disease for prevention of cardiovascular disease and other pathologies in selected patients.(7)

This research will enable us to know the prevalence of metabolic syndrome and its associated factors in T II DM patients in our setting. Knowing the magnitude will significantly help to give the appropriate emphasis and it will also help the responsible health personnel’s including physicians. It will also help to know which associated factors are more prevalent in our population and will be an input in that regard. Overall, the significance of this study for the betterment of patient care in our institution as well as for the country is enormous.

## 2. Literature review

### 2.1 Diabetes mellitus burden

Diabetes is a serious, chronic disease that occurs either when the pancreas does not produce enough insulin (a hormone that regulates blood sugar, or glucose), or when the body cannot effectively use the insulin, it produces. Diabetes is an important public health problem, one of four priority noncommunicable diseases (NCDs) targeted for action by world leaders. Both the number of cases and the prevalence of diabetes have been steadily increasing over the past few decades (8).

Type 2 diabetes mellitus (T2DM) is the predominant form of diabetes worldwide, accounting for 90% of cases globally.1,2 An epidemic of T2DM is under way in both developed and developing countries, although the brunt of the disorder is felt disproportionately in non-European populations. In the Pacific Island of Nauru, diabetes was virtually unknown 50 years ago and is now present in approximately 40% of adults. Globally, the number of people with diabetes is expected to rise from the current estimate of 285 million in 2010 to 438 million in 2030, both figures substantially higher than even recent estimates. T2DM has become one of the world’s most important public health problems (9).

Diabetes caused 1.5 million deaths in 2012. Higher-than-optimal blood glucose caused an additional 2.2 million deaths, by increasing the risks of cardiovascular and other diseases. Forty-three percent of these 3.7 million deaths occur before the age of 70 years. The percentage of deaths attributable to high blood glucose or diabetes that occurs prior to age 70 is higher in low- and middle-income countries than in high-income countries. The most of patients with diabetes are affected by type 2 diabetes. This used to occur nearly entirely among adults, but now occurs in children too(8).

Based on research done on Prevalence of Diabetes Mellitus and Associated Factors in Addis Ababa Public Health Facilities, Addis Ababa, Ethiopia in 2016. The overall prevalence of diabetes mellitus was 14.8%, with a sex-specific pre-valence of 18.35% and 16.62% for males and females, respectively. Older age participants had higher risks of developing diabetes mellitus than younger age individuals. Alcohol drinkers had more risks of developing diabetes mellitus than non-alcohol drinkers. Participants with plasma HDL-C ≥40mg/dl were more likely to develop diabetes mellitus than those with <40mg/dl. Participants with a higher level of plasma triglyceride ≥130mg/dl were found to be more exposed to the risks of developing diabetes mellitus than study participants with a low level of triglycerides(10).

Research from Hosana town shows that the overall prevalence of diabetes was found to be 5.7% (95% CI; 4.0–7.7), out of which more than one third (36%) were not aware of it prior to the survey. Nearly two thirds (61.1%) of the diabetic participants were also found to be hypertensive. In the multi-variable analysis, diabetes was associated with current alcohol use, sitting on average of more than 8 hours/day, abnormal BMI and being hypertensive(11).

A community survey done in southern nations dictates that the overall prevalence of diabetes (1.9%) from our survey was so high in rural setting that prevention and control mechanisms should be designed. The risk factors include advanced age (β=1.04(0.57-1.50)), systolic hypertension (β=0.59(0.01-1.18)), high waist circumference (β=0.86(0.34-1.39)) and postsecondary education (β=0.87(0.03-1.71))(12).

### 2.2 Metabolic syndrome burden

The prevalence of metabolic syndrome in India is observed to be 20.7%. The study reflected significant association with age (p=0.001), gender (p<0.001), waist circumference (p<0.001), BMI (p<0.001), hypertension (p<0.001), FPG (p<0.001) and dyslipidemia (p<0.05). This study revealed prevalence of Metabolic Syndrome to be 20.7% in the study population and outlined the risk factors associated with it. This study would aid in formulating strategies for lifestyle modification, behavioral therapy, diet education and nutritional therapy(13).

According to a systematic review and metanalysis done in Sub-Saharan African countries, the overall prevalence of MS according to the different diagnostic criteria was: IDF: 18.0% (95%CI:13.3-23.3), IDF-ethnic: 16.0% (95%CI:11.3-21.4), JIS: 23.9% (95%CI: 16.5-32.3), NCEP-ATP III: 17.1% (95%CI:12.8-22.0) and WHO: 11.1% (95%CI:5.3-18.9). The prevalence of Metabolic syndrome was higher in women than in men, and higher in (semi-)urban than in rural participants. The Metabolic syndrome prevalence was highest in Southern Africa, followed by Eastern, Western and Central Africa(14).

Research article which was done in Northern Ethiopia on Magnitude, components and predictors of metabolic syndrome, the study revealed that unadjusted and adjusted prevalence rate of Metabolic Syndrome (Metabolic Syndrome) were (CPR = 33.79%; 95%CI: 31.29%–36.36%) and (APR = 34.2%; 95% CI: 30.31%–38.06%) respectively. The most prevalent Metabolic Syndrome component was low HDL concentration (CPR = 70.91%; 95%CI: 68.47%–73.27%) and (APR = 70.61; 95%CI; 67.17–74.05). While high fasting blood glucose (CPR = 20.01% (95%CI: 18.03–22.12) and (APR = 21.72; 95%CI; 18.41–25.03) was the least ones. Eating vegetables four days a week, (AOR = 3.69, 95%CI; 1.33–10.22), a salt sauce added in the food sometimes (AOR = 5.06, 95%CI; 2.07–12.34), overweight (AOR = 24.28, 95%CI; 10.08–58.47] and obesity (AOR = 38.81; 12.20–111.04) had strong association with Metabolic Syndrome (15).

### 2.3 prevalence of metabolic syndrome in type II DM Patients

An article on the prevalence of metabolic syndrome in diabetes patients done in Napal showed prevalence of metabolic syndrome as per NCEP/ATP III and IDF criteria to be 71% and 82% respectively. A higher number of cases with metabolic syndrome were detected using IDF criteria as opposed to NCEP/ATP III criteria in diabetes patients. Nepalese diabetes subjects showed high prevalence of abdominal obesity followed by hypertension(3).

The prevalence of Metabolic Syndrome was 58% in the studied Ghanaian population. Hypertension was the commonest risk factor (60%), followed by central obesity (48.67%) and dyslipidemia (37%). Female type 2 diabetics had a higher prevalence of Metabolic Syndrome and carried more components than their male counterparts. Regression analysis showed three factors; femininity, high body mass index and low educational status were the most critical predictive risk factors of Metabolic Syndrome, according to this study(16).

In another study done in Ghana at Komfo Anokye teaching Hospital the prevalence of Metabolic syndrome was found to be 90.6%. Compared to the above mentioned study which was done in the same country Ghana the prevalence of Metabolic syndrome in type II DM patients is much higher this shows the prevalence could vary in the same country even though in later study the most of the participants were females(17).

In a meta-analysis done in Sub-Saharan African, a total of 23 studies with 6,482 study participants were included. The estimated prevalence of metabolic syndrome in Sub-Saharan African countries was 59.62% (95% CI: 52.20, 67.03). Based on the subgroup analysis, the highest prevalence of metabolic syndrome (61.14%, 95% CI: 51.74, 70.53) was reported in Ethiopia. Additionally, the highest prevalence of metabolic syndrome was reported across studies using the diagnostic criteria of National Cholesterol Education Program Adult Treatment Panel III 64.8% (95% CI: 54.74, 74.86), followed by International Diabetic Federation (57.15%), and World health Organization (53.12%) definitions(18).

Another study done in Sub-Saharan African, they collected clinical and biochemical data for 308 patients with type 2 diabetes (men 157) at the National Obesity Center of the Yaoundé Central Hospital, Cameroon. Concordance was assessed with the use of the Kappa statistic(19).

A study done in Sub-Saharan Africa comparing magnitude of metabolic syndrome base on either IDF or NCEP-ATP criteria showed the prevalence of Metabolic syndrome to be 71.7% according to the IDF criteria and 60.4% according to NCEP-ATP III criteria. The prevalence was significantly higher in women than in men independently of the criteria used (both p< 0.001). Overall concordance between both definitions was low to average 0.51 (95% confidence interval: 0.41–0.61). Combining the two sets of criteria marginally improved the yield beyond that provided by the IDF criteria alone in men, but not in the overall population and in women(19).

Study done in Dessie Referral Hospital shows the proportion of metabolic syndrome was 50.3%, 59.4% and 64.5% according to 2005 International Diabetes Federation, revised ATP III and 2009 harmonized criteria, respectively. Being female (AOR = 2.43; 95% CI = 1.40, 4.21), consumption of red meat (AOR = 2.61; 95% CI = 1.28, 5.33), sedentary leisure time activity (AOR = 2.65; 95% CI = 1.47, 4.78), coffee intake (AOR = 0.43; 95% CI = 0.21, 0.86), BMI 25 kg/m2 (AOR = 9.59; 95% CI = 4.98, 18.47), 40–49 years of age (AOR = 2.74, 95% CI = (1.02, 7.37), 50–59 years of age (AOR = 4.22; 95% CI = 1.60, 11.11) and 70 years of age (AOR = 4.51, 95% CI = 1.44, 14.15) were significantly associated with metabolic syndrome(20).

A cross-sectional study was conducted at Hawassa university comprehensive specialized hospital. The study involved 314 study participants selected by simple random sampling technique. The prevalence of metabolic syndrome according to IDF, WHO, harmonized, and NCEP-ATP III diagnostic criteria was 59.9%, 31.2%, 65.6%, and 70.1%, respectively. According to this study the maximum agreement between IDF and NCEP criteria (K=0.54, P<0.001) and IDF and Harmonized was (K=0.65, P<0.001). Uric acid level was associated factor of metabolic syndrome by all the four definitions, and total cholesterol was associated factors by the three definitions(21).

Another cross-sectional study was conducted at the Diabetes Clinic of the Gondar Hospital and Metabolic Syndrome prevalence was estimated using NCEP ATPIII and IDF criteria. Anthropometric measurements, investigations of serum glucose and lipid profiles were done. A total of 159 participants were included in the study; 119 (59.7%) were females with mean (±SD) age of (49.8±8.7) year. The prevalence of Metabolic Syndrome was 66.7% in NCEP-ATP III and 53.5% in IDF definitions. The most prevalent component of Metabolic Syndrome was elevated triglyceride (56.6% in ATPIII and 62.3% in IDF criteria), followed by abdominal obesity (61%) IDF and elevated blood pressure (55.4%) NCEP-ATPIII criteria. The regression analysis showed that increased age, being female, high BMI, having diabetes for over 5 years and poor glycemic control were significantly associated with metabolic syndrome(17).

### 2.4 Conceptual framework

**Figure 1 2.**
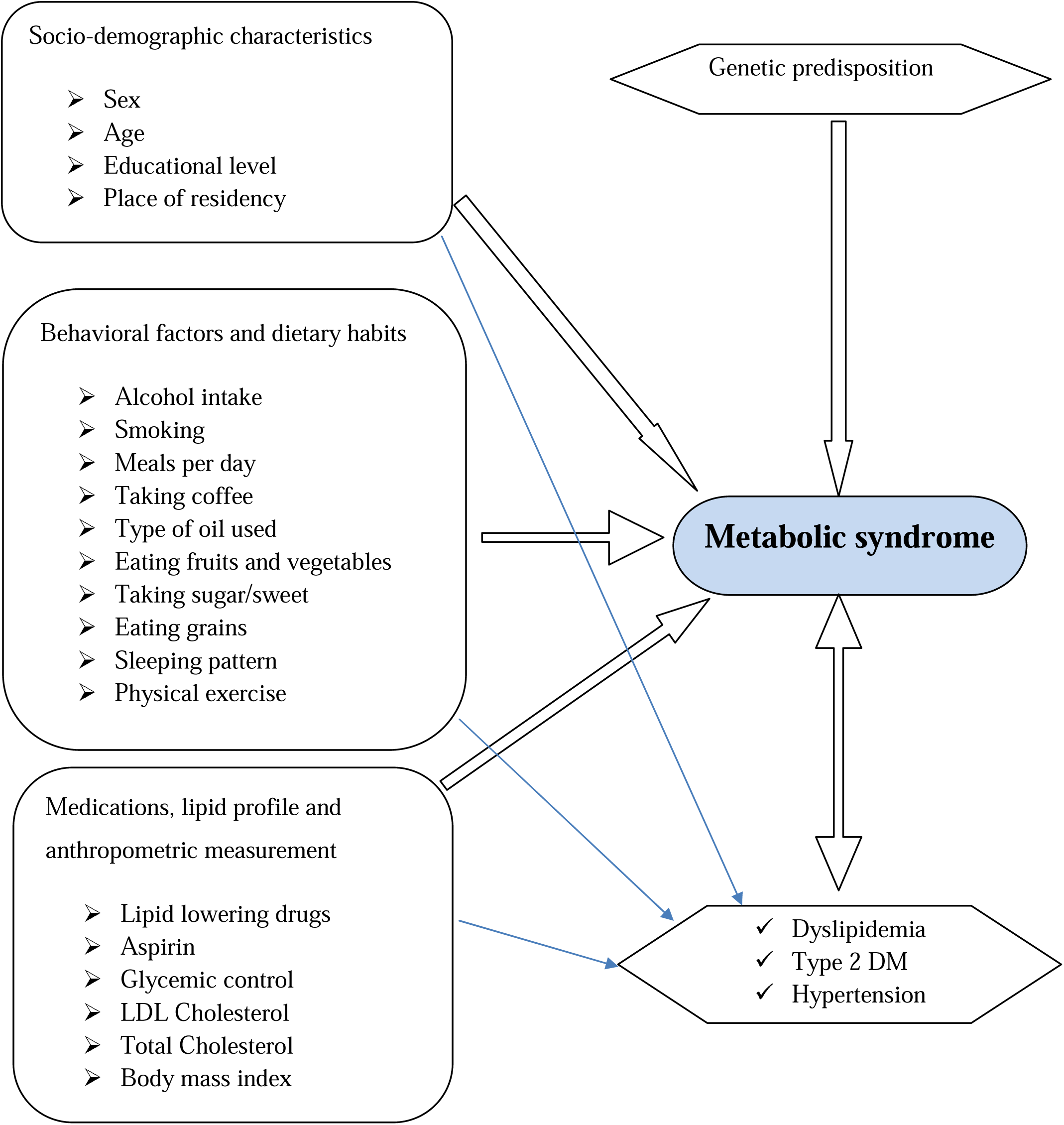
Conceptual framework showing effect of comorbidities and associated factors on Metabolic syndrome.

## 3. Objective

### 3.1. General objective

➢ To determine the Magnitude of metabolic syndrome and its associated factors among patients with type 2 diabetes mellitus who are on follow up at Asella Referral and Teaching Hospital, Diabetes follow up clinic.

### 3.2. Specific objectives

➢ To determine the prevalence of each component of Metabolic Syndrome in T II DM patients.
➢ To assess risks of Metabolic syndrome.
➢ To assess the trend of using statins and aspirin for type 2 DM patients.

## 4. Methods and materials

### 4.1 Study Area

The study was conducted in Asella Hospital which was founded in 1964; renovated in 1992 by Ethio-Italian cooperation and staring from 2014 it is teaching hospital for the newly founded Arsi University, situated in Asella Town, 175 km South-East of Addis Ababa. The hospital has 321 beds and acts as a medical referral center for a population of 3.5 million inhabitants in the Arsi Zone and its surroundings. The outpatient service includes a number of clinics from each clinical department. Under the department of Internal Medicine, there are several medical outpatient clinics and senior referral follow-up clinics. The medical follow-up had two main parts. DM clinic works whole working day and another senior medical follow up clinic works three days per week and over 230 patients are seen every week. Each follow-up clinic had 3 nursing staffs, 2 senior residents and at least 1 senior each day with subspecialists at a time when they are available(22).

### 4.2 Study period

This study was conducted from June 1, 2022, to November 30: 2022 G.C

### 4.3 Study Design

Cross-sectional, prospective, hospital-based survey was conducted to assess the prevalence of metabolic syndrome and its associated factors in type II DM patients who visited diabetic follow-up clinic in ARTH, Ethiopia.

### 4.4 Population

#### 4.4.1 Source population

All patients who had regular follow-up at ARTH, diabetic follow-up clinic was evaluated.

#### 4.4.2 Study population

All type II DM patients who visited ARTH, diabetic follow-up clinic in the study period was included in the study.

### 4.5 Inclusion and Exclusion Criteria

#### 4.5.1 Inclusion Criteria

Patients who meet all the following criteria and are willing to participate in this study was candidates for recruitment:

➢ All physician diagnosed type II DM patients.

#### 4.5.2 Exclusion Criteria

Patients who meet the following criteria was excluded from participation.

➢ All type I DM patients who have follow-up at the clinic and those age below 30 years.

### 4.6. Sample size and sampling technique

#### 4.6.1 Sample size

#### The sample size was determined using a single population proportion formula

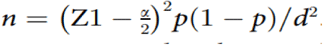

Where:

n = sample size,

p = proportion of Metabolic Syndrome among T2DM patients in Hawassa university comprehensive specialized hospital (71%) (21).

Z1-α/2 = standard normal deviation at 95% confidence interval (1.96), and d = margin of error (5%).

The calculated sample size was 316.

Since the number of total population expected is < 10,000 (N < 10,000) the size of sample will adjusted by the following formula:

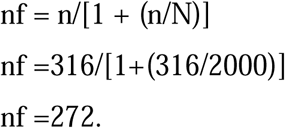

(Total expected patients come to follow up on the study time is 2000).

After considering 5% non-response rate, a final sample size became **285**.

#### 4.6.2 Sampling technique

Systemic random sampling technique was applied after having the list of monthly patient flow to the follow up clinic. Most patients had appointment monthly follow-up (1100 patients/month) and about 50 patients visit diabetic clinic on daily basis. The first patient was selected randomly and every tenth patient on the list were chosen as a participant.

### 4.7. Operational definition

**Dietary Diversity Score (DDS).** Individuals DDS were grouped into low (<4 food groups), medium (4 to 6 food groups), and high (7 to 9 food groups)(23).

**Knowledge.** Seven multiple-choice questions about Metabolic Syndrome with a total of 17 responses (Cronbach’s alpha = 0.82) will be asked and summed. Then individuals’ knowledge will be grouped into low (<51%), fair (51 to 80%), and good (*>*80%)(24).

**Physical activity.** Individuals who did moderate-intensity activity for at least 30 minutes per day on at least five days per week were considered as physically active(25).

**Adequate sleep.** Participants who sleep 7 to 8 hours per day without difficulty initiating and maintaining sleep are grouped as having adequate sleep(26).

**Metabolic syndrome.** Three definitions of Metabolic Syndrome were considered. However, a definition that had better agreement with the rest two definitions, after calculating Kappa statistics, was used in this study to run the regression analysis.

**The Revised National Cholesterol Education Program: Third adult treatment panel (ATP III) definition.** Metabolic Syndrome is the group of at least 3 of the following 5 risk factors: WC *>*102 for men and *>*88 cm for women, BP *>*130/85 mmHg or on treatment, FBG > 110 mg/dl or on treatment, TG >150 mg/dl or on treatment and HDL *<* 40mg/dl for men and *<* 50mg/dl for women(2).

**International Diabetes Federation (IDF) definition.** Metabolic Syndrome is WC *>* 80cm for women, *>* 94 cm for men plus 2 or more the factors (BP > 130/85 mmHg or on treatment, TG > 150 mg/dl or on treatment HDL *<* 40 mg/dl for men and *<* 50 mg/dl for women or on treatment, and FBG > 100 mg/dl)(4).

**The 2009 harmonized criteria.** Metabolic Syndrome is a cluster of three or more of the 5 interrelated risk factors (TG > 150 mg/dl, HDL *<* 40 mg/dl for men and *<* 50mg/dl for women, BP *>* 130/85 mmHg, FBG > 100 mg/dl, and WC *>*102 cm for men and *>*88 cm for women)(27).

**Body mass index (BMI).** It is classified into underweight (*<*18.5 kilogram per meter square (kg/m2)), normal weight (18.5–24.9kg/m2), overweight (25–29.9 kg/m2) and obese (*>*30 kg/m2)(28).

### 4.8. Study Variables

The following variables was measured.

#### 4.8.1. Dependent variables

➢ Metabolic Syndrome

#### 4.8.2. Independent variable

­ Demographic characteristics: Age, Sex, Residency, Marital status, Occupation, Income, Educational status, Lifestyle.

➢ Habit of doing exercise
➢ Dietary habit
➢ Use of Additional drugs for comorbidities
➢ Current and Past smoking
➢ Knowledge about Metabolic Syndrome
➢ Sleep pattern
➢ Presence of psychological stress
➢ BMI

### 4.9. Data collection tools and measurements

Data was collected by the World Health Organization stepwise approach for non-communicable disease surveillance (25). Data about socio-demographic, medication, behavioral, and dietary factors will be collected by a structured questionnaire. Height was measured in a standing position by a height measuring board on barefoot. Weight was measured using The Swiss Private Equity & Corporate Finance Association (SECA) weight measuring scale. WC was measured by a normal tension measuring tape at the midpoint of the inferior margin of the last rib and the iliac crest at the end of expiration. Each anthropometric measurement was recorded to the nearest 0.1cm (kg for weight). Blood pressure was measured by sphygmomanometer from the right arm after participants rested for 5 minutes. Data was collected by 4 trained BSc nurses.

FBG was revied from patients’ chart since it will be done on every visit. lipid profiles were determined from serum blood using Auto chemistry analyzer by laboratory technologists following the standard operating procedures. Additionally, the intake of medication was assessed from the patient card.

### 4.10. Data collectors

Data was collected by total of four BSC nurses after they are trained for half day about the study matter. They were also given training on objective of the study, variables on the questionnaire and its implication. Then, they were assigned to fill the data collection format. All data collection activities were supervised by principal investigator.

### 4.11. Data analysis, processing, and Interpretation

Collected data was first cleaned, edited and entered into a computer and analyzed using latest version of software program SPSS. A P-value of <0.05 was considered statistically significant.

Then, the findings were discussed in detail, and compared with similar studies done elsewhere. Based on the results, recommendations and conclusions was made and the document would be disseminated.

### 4.12. Data quality assurance

Pre-testing of data collection tools was made to ensure data quality. Adequate training was provided for data collectors, and the compilation format was prepared in simple English to maintain clarity and easier understanding by those data collectors. The collected data was then be checked for completeness and consistency on the day of collection by the principal investigator and any inconsistency, inaccuracy, or missing data implied was returned for correction on same day. There was demonstration and a practical session on interviewing and record reviewing with the data collectors. Onsite supervision was done by principal investigator.

### 4.13. Ethical consideration

Ethical clearance to conduct the study was obtained from the ethical Review Board of Arsi University before official commencement of the data collection process. A letter of recommendation was also obtained from the responsible office to the head of out-patient clinic and internal medicine ward. Written informed consent was obtained from all patients before data collection. Patient’s confidentiality, equity of services and interests of patients was ensured during the study period. All patients participating in this study were offered clinical care equally with other patients.

### 4.14. Dissemination of the results

After research completion and finalizing the report, the findings of the study will be disseminated to all relevant stakeholders through presentation and publication in peer-reviewed reputable journal. Copies of the research will be given to Arsi University College of Health Science, to the department of Internal Medicine, to the ministry of health and Ethiopian Non-communicable diseases association, other concerned institutions, and stake holders for possible intervention based on the findings.

## 5. Results

### 5.1 Socio demographic characteristics of the study population

A total of 285 Type II DM patients were enrolled in this study. The number of males in this study was 178 (62.5%) and females was 107 (37.5%). The age of the study participants were 4 (1.4%) less than 30 years, 7 (2.5%) between 30-39, 81 (28.4%) between 40-49, 76 (26.7%) between 50-59, 92 (32.3%) between 60-69 and 25 (8.8%) above 70 years Figure 5 1. 215 (75.4%) of them at least completed primary education and 77 (27%) of them have diploma or above. Most of the participants were married 261 (91.6%), the rest are single (0.4%), divorced (4.2%) and widowed (3.9%). Concerning their occupational status 66 (23.2%) were Housewife, 33 (11.6%) were Government employed, 56 (19.6%) were Farmers, 33 (11.6%) were Marchant, 51 (17.9%) were retired, and 46 (16.1%) had private job. Participants living in urban were 198 (69.5%) Table 5 1

#### 5.1.1 Age of participants

**Figure 5 1.**
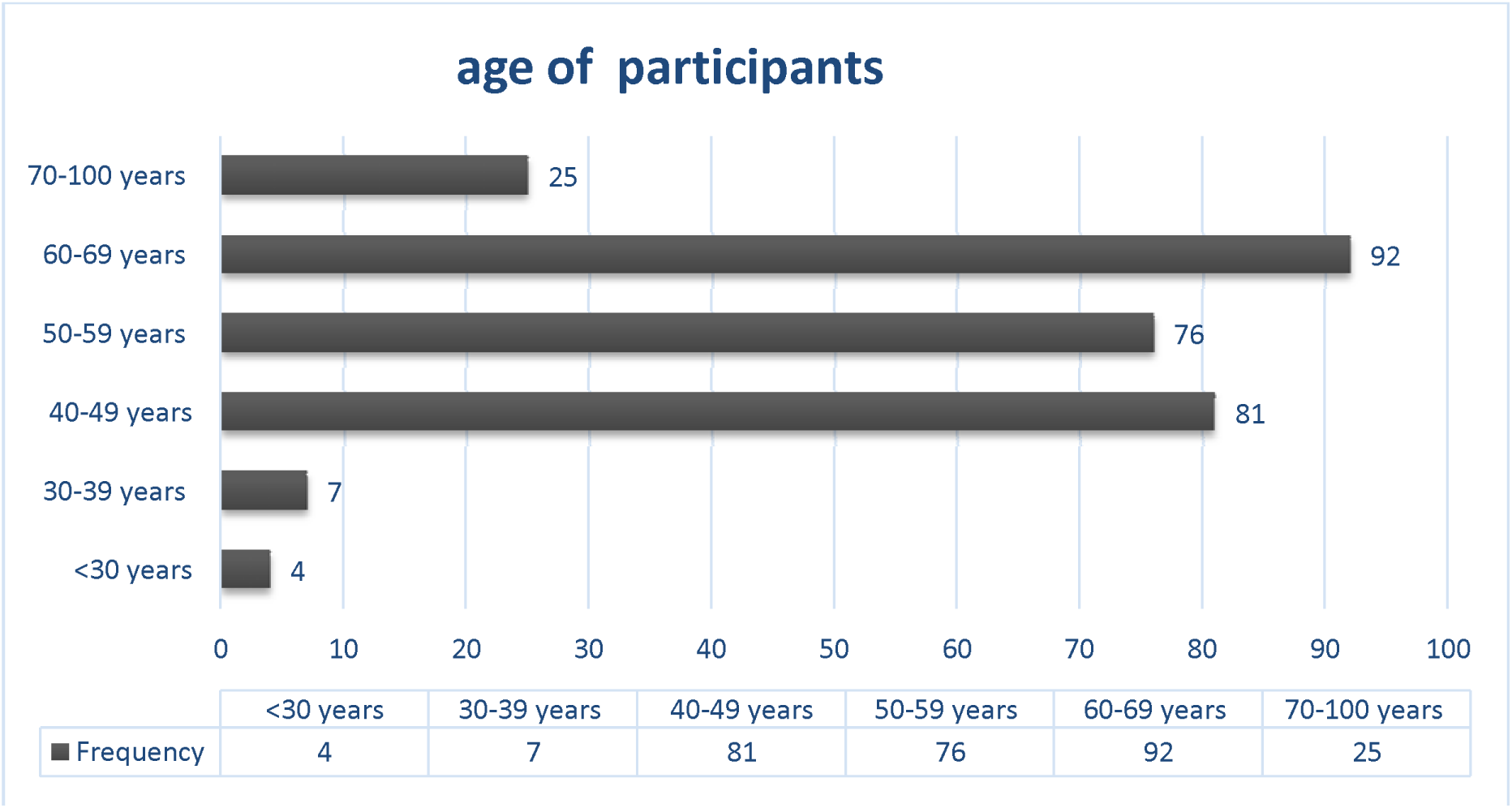
Age of participants at ARTH, Oromia, Ethiopia from July to September 2022 EC. n(285)

**Table 5 1.** Socio demographic characteristics of the participants at ARTH, Oromia, Ethiopia from July to September 2022 EC. n(285)

| *Variable* | *Category* | *Frequency* | *Percent* |
|---|---|---|---|
| *Sex* | Male | 178 | 62.5 |
|  | Female | 107 | 37.5 |
| *Age group in years* | <30 | 4 | 1.4 |
|  | 30-39 years | 7 | 2.5 |
|  | 40-49 years | 81 | 28.4 |
|  | 50-59 years | 76 | 26.7 |
|  | 60-69 years | 92 | 32.3 |
|  | 70-100 | 25 | 8.8 |
| *Educational level* | cannot read and write | 70 | 24.6 |
|  | primary education | 86 | 30.2 |
|  | secondary education | 52 | 18.2 |
|  | diploma and above | 77 | 27.0 |
| *Marital status* | single | 1 | 0.4 |
|  | married | 261 | 91.6 |
|  | divorced | 12 | 4.2 |
|  | widowed | 11 | 3.9 |
| *Occupational status* | Housewife | 66 | 23.2 |
|  | Government employed | 33 | 11.6 |
|  | Farmer | 56 | 19.6 |
|  | Merchant | 33 | 11.6 |
|  | Retired | 51 | 17.9 |
|  | Other private job | 46 | 16.1 |
| *Place of residency* | Urban | 198 | 69.5 |
|  | Rural | 87 | 30.5 |

### 5.2 Medications and behavioral factors

Of the total study participants one hundred eighty-eight (66%) had Diabetes for more than five years. Hundred thirty-nine (48.8%) of them were taking oral antidiabetic medications, eighty-two (28.8%) insulin and sixty-four (22.5%) of them were taking both oral antidiabetic medication and insulin Figure 5 2. 133 (46.7%) had the diagnosis of hypertension and 158 (55.4%) were taking lipid lowering medications. Study participants who were taking Aspirin were 58 (20.4%) Figure 5 3. Among them only one participant was active cigarette smoker and 7 of them were former smokers. Nine participants chew khat, 110 (38.6%) do regular physical activity and only 52 (18.5%) sleep for less than eight hours a day Table 5 2.

**Table 5 2.** Medications and behavioral factors of participants at ARTH, Oromia, Ethiopia from July to September 2022 EC. n(285)

| Variable | Category | Frequency | Percent |
|---|---|---|---|
| **Type of medication used** | Tablet | 139 | 48.8 |
|  | Insulin | 82 | 28.8 |
|  | Both | 64 | 22.5 |
| **Duration of DM** | >/=5 years | 188 | 66.0 |
|  | <5 years | 97 | 34.0 |
| **Treatment for Hypertension** | Yes | 133 | 46.7 |
|  | No | 152 | 53.3 |
| **Lipid lowering medication** | Yes | 158 | 55.4 |
|  | No | 127 | 44.6 |
| **Taking Aspirin** | Yes | 58 | 20.4 |
|  | No | 227 | 79.6 |
| **Smoking** | Current smoker | 1 | 0.4 |
|  | Former smoker | 7 | 2.5 |
|  | Never smoked | 277 | 97.2 |
| **Khat chewing** | Yes | 9 | 3.2 |
|  | No | 276 | 96.8 |
| **Regular physical activity** | Yes | 110 | 38.6 |
|  | No | 175 | 61.4 |
| **Sleep pattern** | >/= 8 hours | 233 | 81.8 |
|  | <8 hours | 52 | 18.2 |

#### 5.2.1 Type of medication used for DM

**Figure 5 2.**
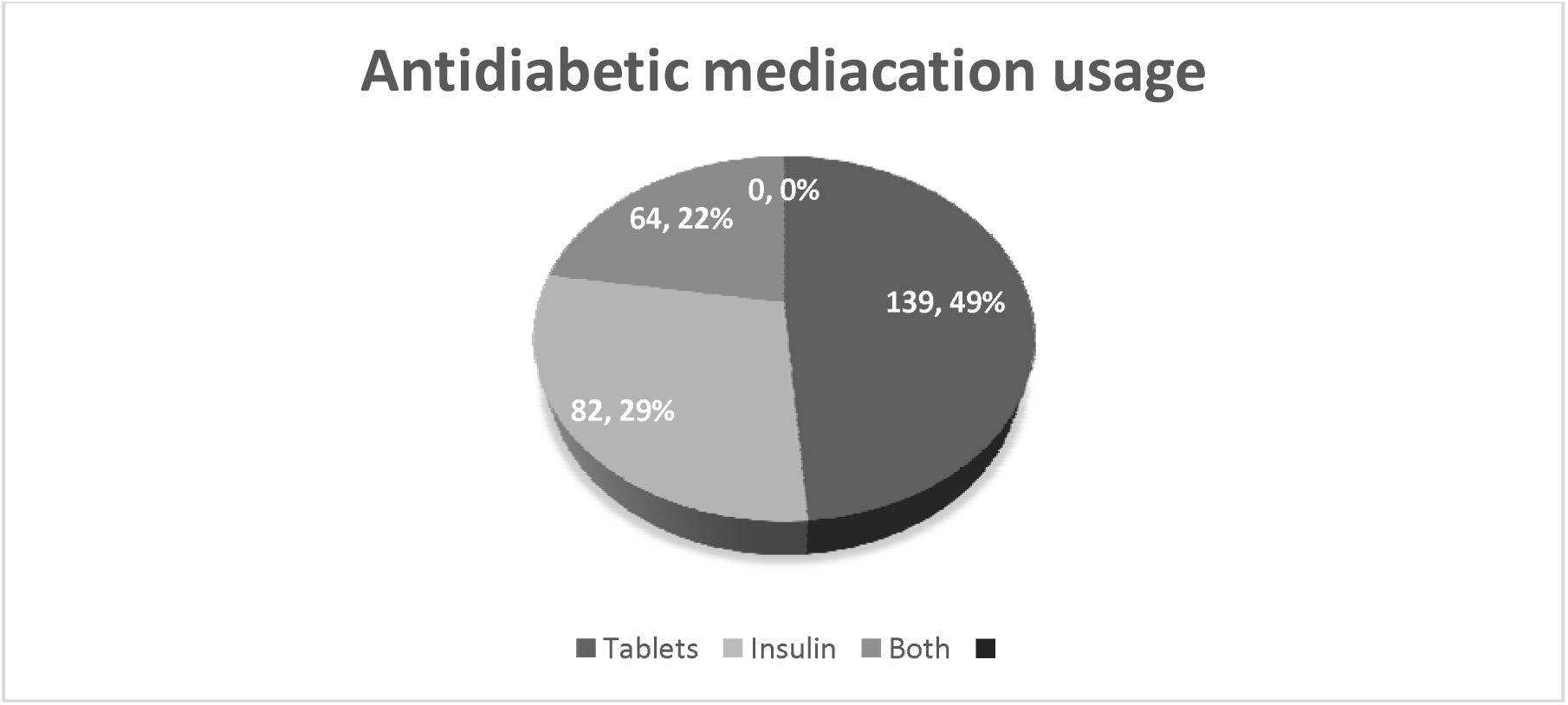
Type of medication used for DM among participants at ARTH, Oromia, Ethiopia from July to September 2022 EC. n(285)

#### 5.2.2 Hypertension, Lipid lowering medication and Aspirin utilization

**Figure 5 3.**
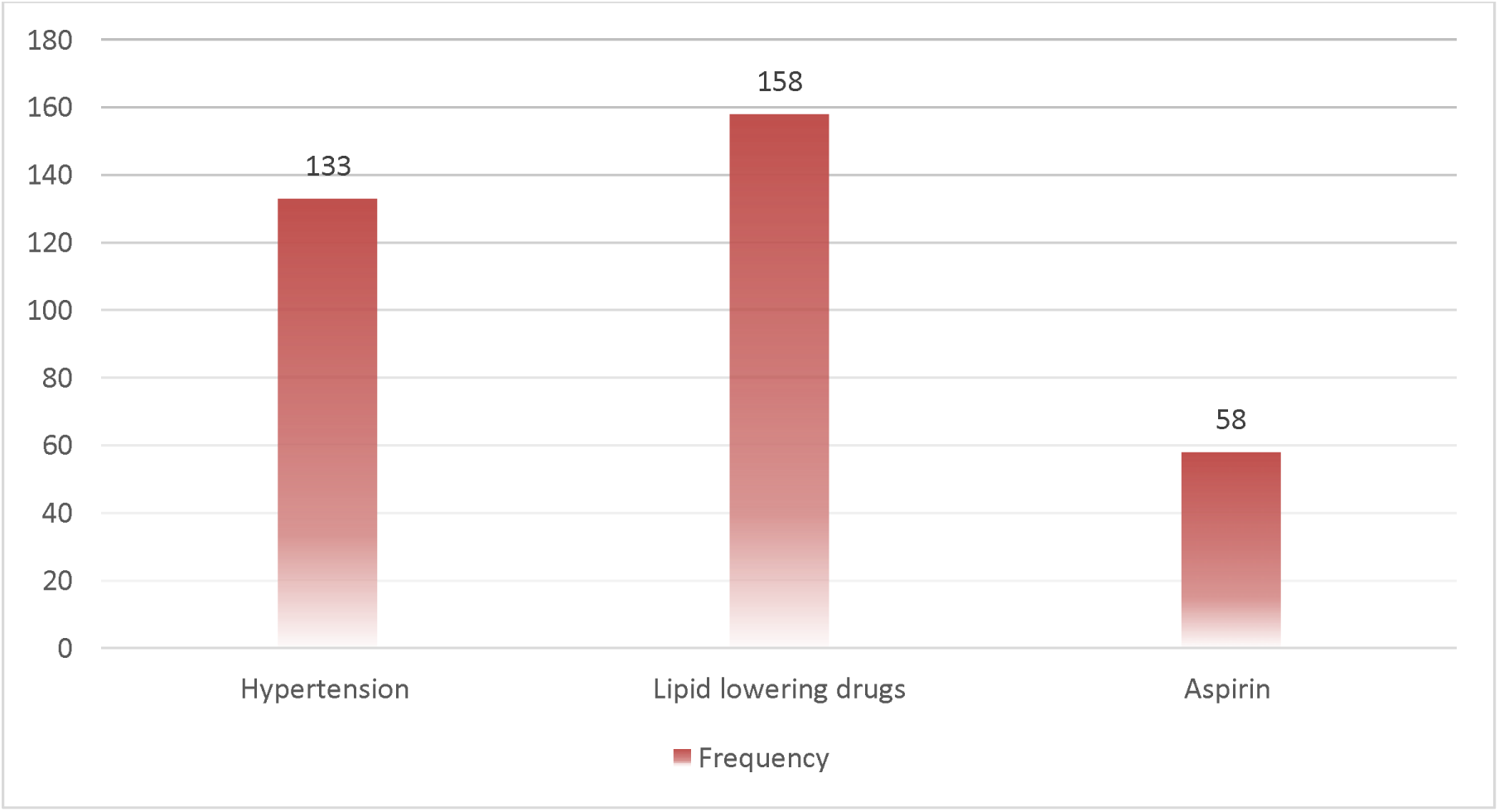
Hypertension, Lipid lowering medication and Aspirin utilization among participants at ARTH, Oromia, Ethiopia from July to September 2022 EC. n(285)

### 5.3 Dietary habits

The dietary habit of study participants including frequency of taking vegetables, fruits, meat, coffee, milk, milk products, processed grains, sugar/sweet and alcohol are depicted below Table 5 3 Dietary habits and their frequency.

**Table 5 3.**
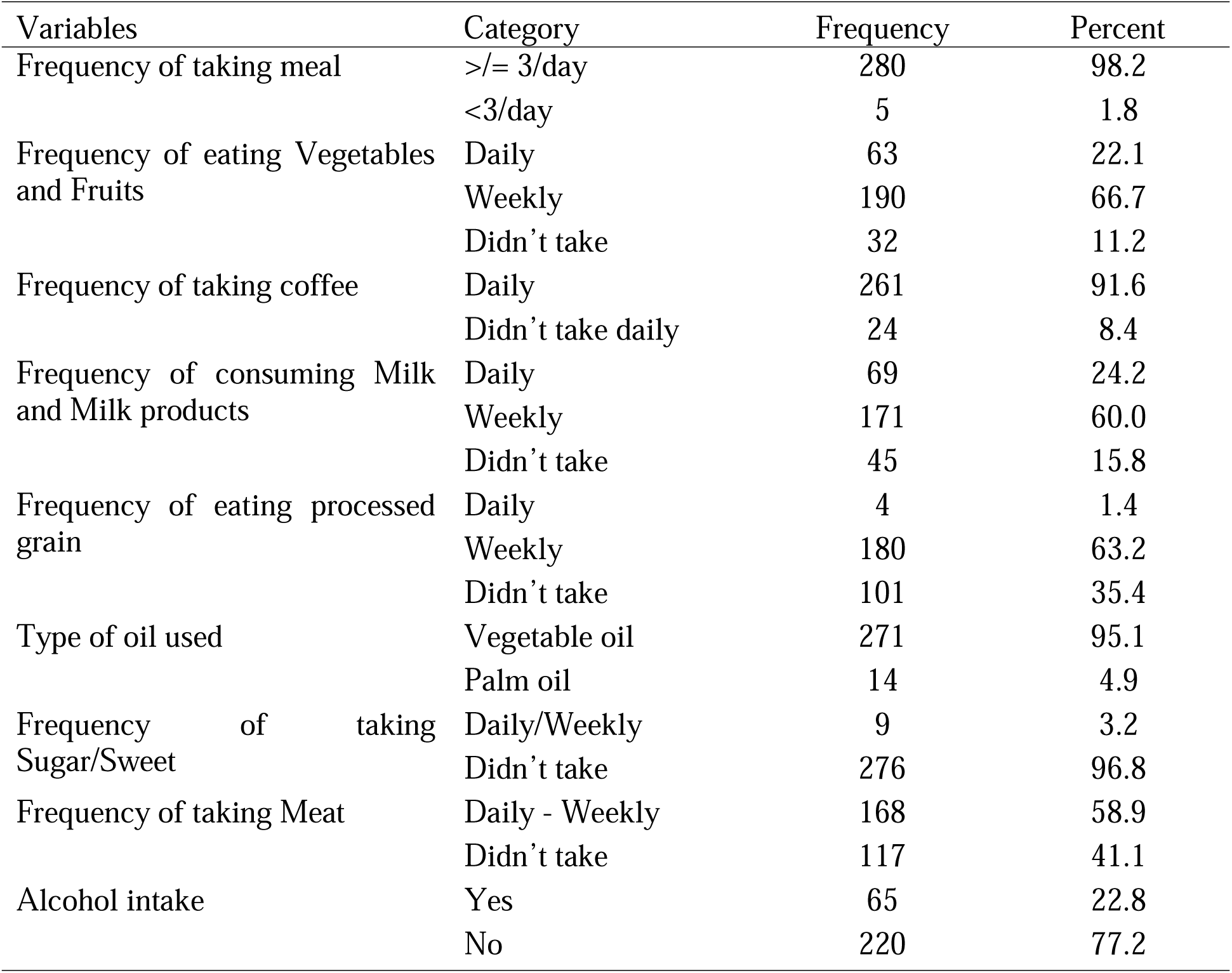
Dietary habits and their frequency of participants at ARTH, Oromia, Ethiopia from July to September 2022 EC. n(285)

### 5.4 Lipid profile, Anthropometric measurement and Clinical characteristics

The minimum and maximum weight of the participants was 42kg and 113kg respectively making the mean 72.77kg and standard deviation 11.97. Among them the minimum and maximum height was 120cm and 212cm respectively, the mean and standard deviation is 166.94 cm and 9 respectively Table 5 5. 124 (43.5%) were overweight, 36 (12.6%) were obese, 8 (2.8%) were underweight and 117 (41.1%) had normal BMI. 159 (55.8%) of the participants had FBS greater than 130mg/dl. Participants who had blood pressure ≥130/85mmhgwere 202 (70.9%). Only 36 (12.6%) of study participants had waist circumference ≥102cm for Men and ≥88cm for females. About 63 (22.1%) and 87 (30.5%) of participants had total cholesterol ≥200mg/dl and LDL ≥130mg/dl respectively. In this study 201 (70.5%) had HDL less than 40mg/dl and 50mg/dl for men and women respectively in other way 131 (46%) of them dad triglyceride more than 150mg/dg Table 5 4.

**Table 5 4.**
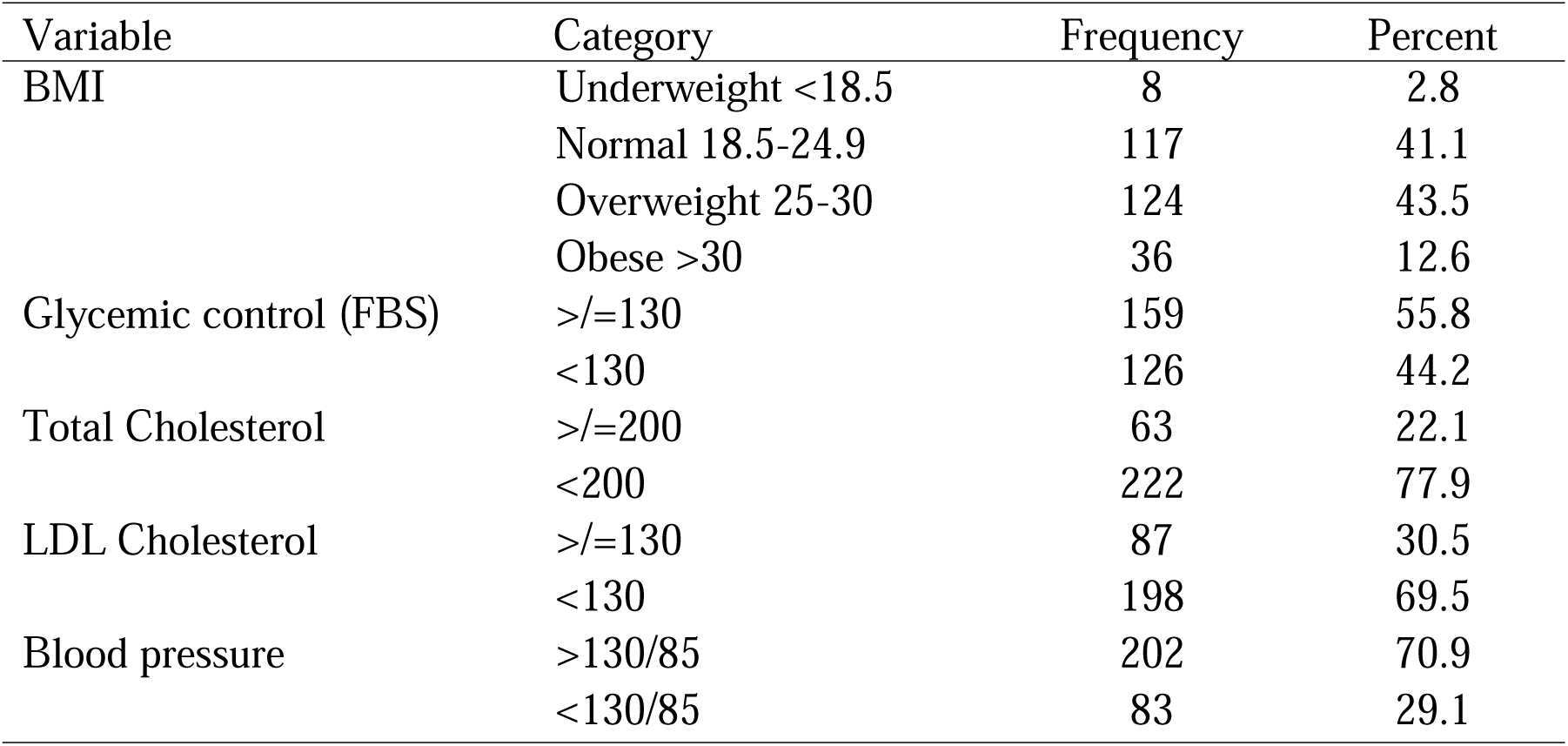
Lipid profile, Anthropometric measurement and Clinical characteristics of participants at ARTH, Oromia, Ethiopia from July to September 2022 EC. n(285)

| Variable | Category | Frequency | Percent |
|---|---|---|---|
| BMI | Underweight <18.5 | 8 | 2.8 |
|  | Normal 18.5-24.9 | 117 | 41.1 |
|  | Overweight 25-30 | 124 | 43.5 |
|  | Obese >30 | 36 | 12.6 |
| Glycemic control (FBS) | >/=130 | 159 | 55.8 |
|  | <130 | 126 | 44.2 |
| Total Cholesterol | >/=200 | 63 | 22.1 |
|  | <200 | 222 | 77.9 |
| LDL Cholesterol | >/=130 | 87 | 30.5 |
|  | <130 | 198 | 69.5 |
| Blood pressure | >130/85 | 202 | 70.9 |
|  | <130/85 | 83 | 29.1 |

**Table 5 5.** Weight and Height of participants at ARTH, Oromia, Ethiopia from July to Septembe 2022 EC. n(285)

| Variable | Max | Min | Mean | Std. Deviation |
|---|---|---|---|---|
| Weight | 113 | 42 | 72.77 | 11.972 |
| Height | 212 | 120 | 166.94 | 9.00 |

### 5.5 Magnitude of metabolic syndrome

The magnitude of meta bolic syndrome in this study was 75.1% (214) ‘ Figure 5 4.

**Figure 5 4.**
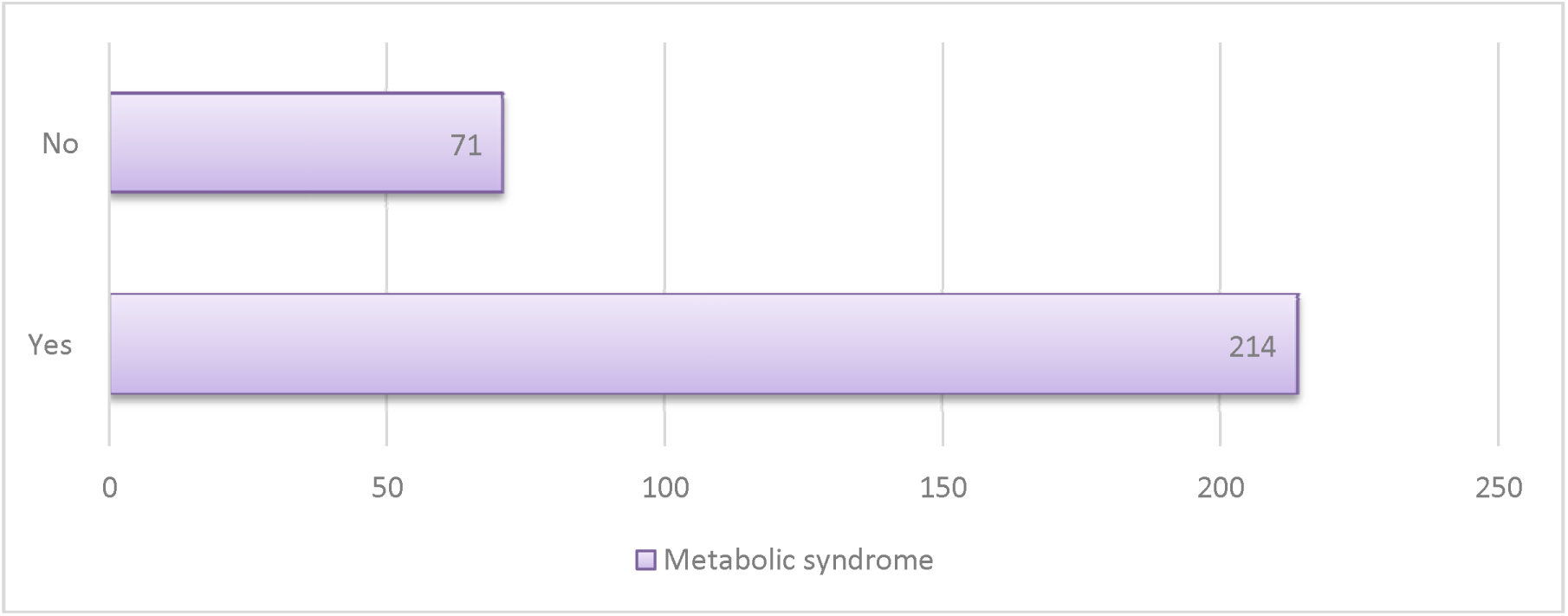
Magnitude of metabolic syndrome of participants at ARTH, Oromia, Ethiopia from July to September 2022 EC. n(285)

#### 5.5.1 Prevalence of Metabolic syndrome among different Socio demographic characteristics

The distribution among different sex was almost comparable 75.3% in male participants and 74.8% in female participants. The prevalence of metabolic syndrome in less than 30 years was 75%, between 30-39 years was 100% between 40-49 years was 67.9%, 50-59 years was 78.9%, 60-69 was 76.1% and above 70 years was 76% Table 5 6.

**Table 5 6.** Prevalence of Metabolic syndrome among different Socio demographic characteristics of participants at ARTH, Oromia, Ethiopia from July to September 2022 EC. n(285)

|  |  | Metabolic syndrome |  |  |  |  |  |
|---|---|---|---|---|---|---|---|
|  |  | No |  | Yes |  | Total |  |
| Variables | Category | Frequency | Percent (%) | Frequency | Percent (%) | Frequency | Percent (%) |
| Sex | Male | 44 | 24.7% | 134 | 75.3% | 178 | 62.5% |
|  | Female | 27 | 25.2% | 80 | 74.8% | 107 | 37.5% |
| Age in years | 0-29 years | 1 | 25.0% | 3 | 75.0% | 4 | 1.4% |
|  | 30-39 years | 0 | 0.0% | 7 | 100.0% | 7 | 2.5% |
|  | 40-49 years | 26 | 32.1% | 55 | 67.9% | 81 | 28.4% |
|  | 50-59 years | 16 | 21.1% | 60 | 78.9% | 76 | 26.7% |
|  | 60-69 years | 22 | 23.9% | 70 | 76.1% | 92 | 32.3% |
|  | 70-100 | 6 | 24.0% | 19 | 76.0% | 25 | 8.8% |
| Educational level | cannot read and write | 13 | 18.8% | 56 | 81.2% | 69 | 24.2% |
|  | primary education | 23 | 26.1% | 65 | 73.9% | 88 | 30.9% |
|  | secondary education | 16 | 31.4% | 35 | 68.6% | 51 | 17.9% |
|  | diploma and above | 19 | 24.7% | 58 | 75.3% | 77 | 27.0% |
| Marital status | single | 0 | 0.0% | 1 | 100.0% | 1 | 0.4% |
|  | married | 65 | 24.9% | 196 | 75.1% | 261 | 91.6% |
|  | divorced | 4 | 33.3% | 8 | 66.7% | 12 | 4.2% |
|  | widowed | 2 | 18.2% | 9 | 81.8% | 11 | 3.9% |
| Occupational status | Housewife | 20 | 30.3% | 46 | 69.7% | 66 | 23.2% |
|  | Government employed | 6 | 18.2% | 27 | 81.8% | 33 | 11.6% |
|  | Farmer | 15 | 26.8% | 41 | 73.2% | 56 | 19.6% |
|  | Merchant | 7 | 21.2% | 26 | 78.8% | 33 | 11.6% |
|  | Retired | 13 | 25.5% | 38 | 74.5% | 51 | 17.9% |
|  | Other private job | 10 | 21.7% | 36 | 78.3% | 46 | 16.1% |
| Place of residency | Urban | 48 | 24.2% | 150 | 75.8% | 198 | 69.5% |
|  | Rural | 23 | 26.4% | 64 | 73.6% | 87 | 30.5% |

Participants who had formal education and at least attended primary education were 80.4%. Place of residency, among 198 participants who lived in Urban area 150 (75.8%) had Metaboli syndrome. From 261 participants who were married 196 (75.1%) had metabolic syndrome Table 5 6.

#### 5.5.2 Prevalence of Metabolic syndrome among different clinical characteristics

**Table 5 7.**
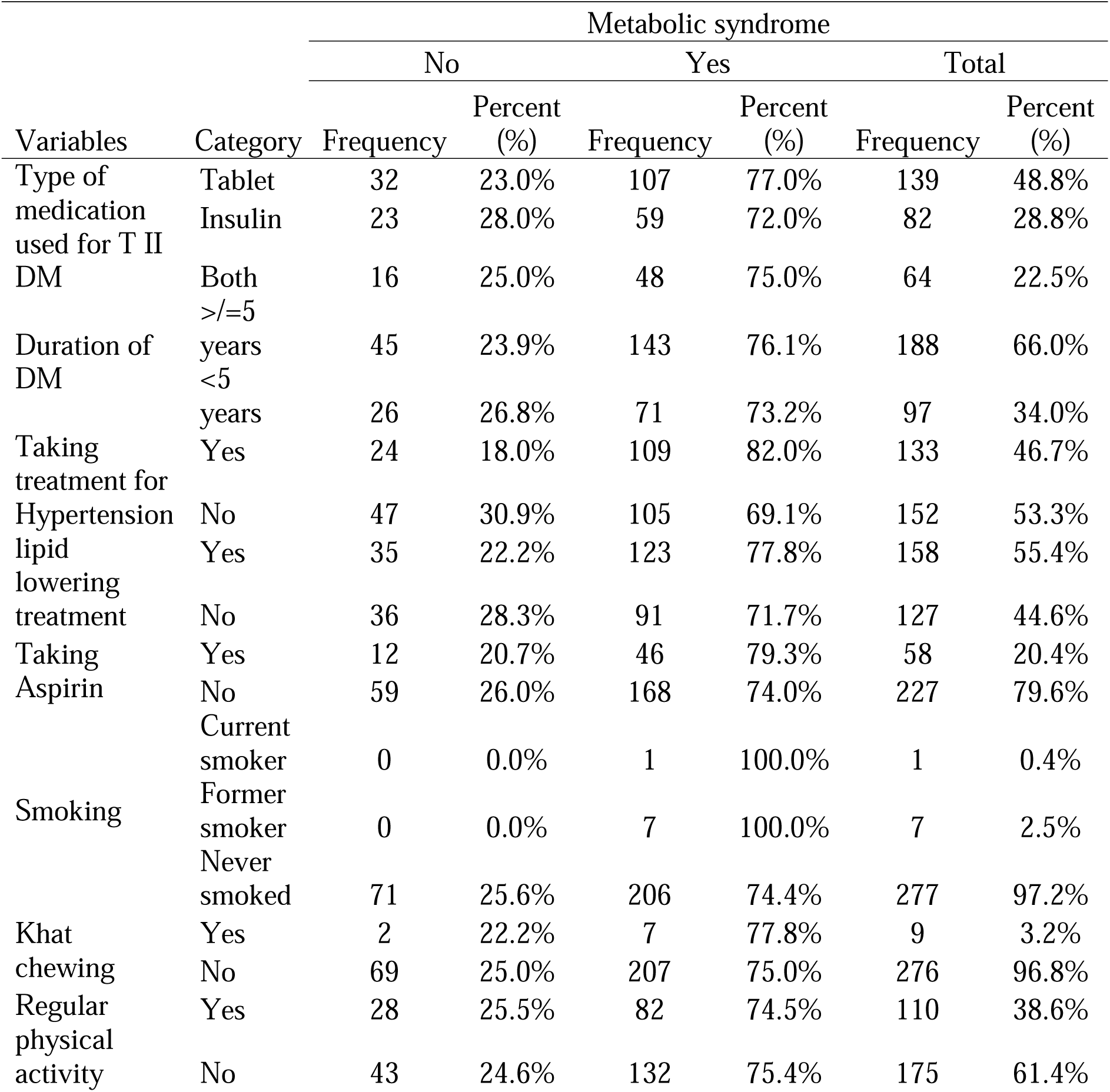
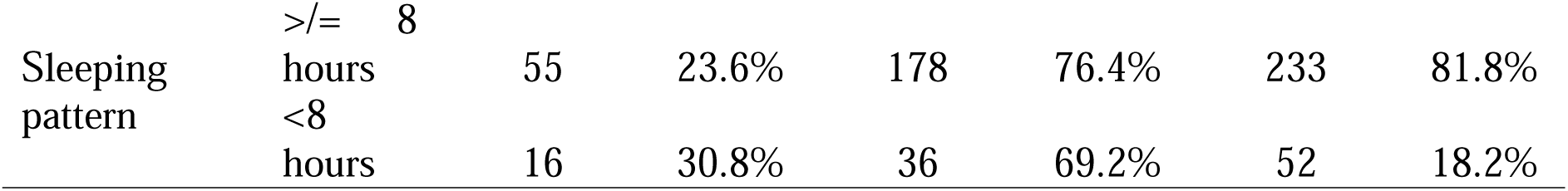
Prevalence of Metabolic syndrome among different clinical characteristics of participants at ARTH, Oromia, Ethiopia from July to September 2022 EC. n(285)

#### 5.5.3 Metabolic syndrome compared to different dietary habits

**Table 5 8.** Metabolic syndrome compared to different dietary habits among participants at ARTH, Oromia, Ethiopia from July to September 2022 EC. n(285)

|  |  | Metabolic syndrome |  |  |  |  |  |
|---|---|---|---|---|---|---|---|
|  |  | No |  | Yes |  | Total |  |
| Variables | Category | Frequency | Percent (%) | Frequency | Percent (%) | Frequency | Percent (%) |
| Frequency of taking per day | >/= 3/day | 70 | 25.0% | 210 | 75.0% | 280 | 98.2% |
|  | <3/day | 1 | 20.0% | 4 | 80.0% | 5 | 1.8% |
| Frequency of eating vegetables and fruits | Daily | 12 | 19.0% | 51 | 81.0% | 63 | 22.1% |
|  | Weekly | 57 | 30.0% | 133 | 70.0% | 190 | 66.7% |
|  | Didn’t take | 2 | 6.3% | 30 | 93.8% | 32 | 11.2% |
| Frequency of taking coffee | Daily | 66 | 25.3% | 195 | 74.7% | 261 | 91.6% |
|  | Didn’t take daily | 5 | 20.8% | 19 | 79.2% | 24 | 8.4% |
| Frequency of consuming milk and milk products | Daily | 15 | 21.7% | 54 | 78.3% | 69 | 24.2% |
|  | Weekly | 49 | 28.7% | 122 | 71.3% | 171 | 60.0% |
|  | Didn’t take | 7 | 15.6% | 38 | 84.4% | 45 | 15.8% |
| Frequency of eating processed grain | Daily | 1 | 25.0% | 3 | 75.0% | 4 | 1.4% |
|  | Weekly | 45 | 25.0% | 135 | 75.0% | 180 | 63.2% |
|  | Didn’t take | 25 | 24.8% | 76 | 75.2% | 101 | 35.4% |
| Type of oil used | Vegetable oil | 68 | 25.1% | 203 | 74.9% | 271 | 95.1% |
|  | Palm oil | 3 | 21.4% | 11 | 78.6% | 14 | 4.9% |
| Frequency of taking sugar/sweet | Daily/Weekly | 1 | 11.1% | 8 | 88.9% | 9 | 3.2% |
|  | Didn’t take | 70 | 25.4% | 206 | 74.6% | 276 | 96.8% |
| Frequency of taking meat | Daily - Weekly | 34 | 20.2% | 134 | 79.8% | 168 | 58.9% |
|  | Didn’t take | 37 | 31.6% | 80 | 68.4% | 117 | 41.1% |
| Alcohol intake | Yes | 8 | 12.3% | 57 | 87.7% | 65 | 22.8% |
|  | NO | 63 | 28.6% | 157 | 71.4% | 220 | 77.2% |

#### 5.5.4 Prevalence of Metabolic syndrome in different Lipid profile, FBS and Anthropometric measurement

**Table 5 9.** Prevalence of Metabolic syndrome in relation to Lipid profile, FBS and Anthropometric measurement among participants at ARTH, Oromia, Ethiopia from July to September 2022 EC. n(285)

|  |  | Metabolic syndrome |  |  |  |  |  |
|---|---|---|---|---|---|---|---|
|  |  | No |  | Yes |  | Total |  |
| Variables | Category | Frequency | Percent (%) | Frequency | Percent (%) | Frequency | Percent (%) |
| BMI (kg/m2) | Underweight <18.5 | 1 | 12.5% | 7 | 87.5% | 8 | 2.8% |
|  | Normal 18.5-24.9 | 38 | 32.5% | 79 | 67.5% | 117 | 41.1% |
|  | Overweight 25-30 | 28 | 22.6% | 96 | 77.4% | 124 | 43.5% |
|  | Obese >30 | 4 | 11.1% | 32 | 88.9% | 36 | 12.6% |
| Glycemic control (FBS) | >/=130 | 35 | 22.0% | 124 | 78.0% | 159 | 55.8% |
|  | <130 | 36 | 28.6% | 90 | 71.4% | 126 | 44.2% |
| Total Cholesterol (mg/dl) | >/=200 | 6 | 9.5% | 57 | 90.5% | 63 | 22.1% |
|  | <200 | 65 | 29.3% | 157 | 70.7% | 222 | 77.9% |
| LDL Cholesterol (mg/dl) | >/=130 | 10 | 11.5% | 77 | 88.5% | 87 | 30.5% |
|  | <130 | 61 | 30.8% | 137 | 69.2% | 198 | 69.5% |

### 5.6 Prevalence of Metabolic syndrome components

Among the criterions of Metabolic syndrome, the most prevalent was Blood pressure ≥130/85 (70.8%) followed by HDL Cholesterol <40 Men and <50 Female (70.5%), Triglyceride ≥150 (46%) and Waist circumference ≥102 Men, ≥88 Female (12.6%) respectively Table 5 10.

Prevalence of each component of Metabolic syndrome according to sex of participants was done. From the components of metabolic syndrome high Waist circumference and Triglyceride level were associated with female sex than male whereas Hypertension occurred more in males. The likelihood of females to have high Waist circumference (≥102 men, ≥88 female) was 1.8 times that for males. The same hold true for Triglyceride level Females have 1.81 times higher Triglyceride level (≥150) compared Males. High blood pressure (Bp≥130/85) occurred 1.63 more likely in males than the female.

**Table 5 10.** Prevalence of Metabolic syndrome components among participants at ARTH, Oromia, Ethiopia from July to September 2022 EC. n(285) Metabolic syndrome.

|  |  | Metabolic syndrome |  |  |  |  |  |
|---|---|---|---|---|---|---|---|
|  |  | No |  | Yes |  | Total |  |
| Variables | Category | Frequency | Percent (%) | Frequency | Percent (%) | Frequency | Percent (%) |
| Blood pressure (mmhg) | >130/85 | 21 | 10.4% | 181 | 89.6% | 202 | 70.9% |
| Waist circumference (cm) | ≥102 men,<br>≥88 female | 2 | 5.6% | 34 | 94.4% | 36 | 12.6% |
| HDL Cholesterol (mg/dl) | <40 men,<br><50 female | 27 | 13.4% | 174 | 86.6% | 201 | 70.5% |
| Triglyceride (mg/dl) | ≥150 | 4 | 3.1% | 127 | 96.9% | 131 | 46.0% |

### 5.7 Utilization of lipid lowering medication and Aspirin

According to American diabetic association at least moderate intensity statins are recommended for type II DM patients age above 40 years. Our study showed that only 158 (55.4%) of participants were taking lipid lowering medication. 58 (20.4%) of the participants was taking Aspirin and according to American diabetic association use of Aspirin is recommended for type II DM patients base on ASCVD risk score (7).

**Table 5 11.** Utilization of lipid lowering medication and Aspirin among participants at ARTH, Oromia, Ethiopia from July to September 2022 EC. n(285)

| Variables | Category | Frequency | Percentage % |
|---|---|---|---|
| Taking lipid lowering medications | Yes | 158 | 55.4% |
|  | No | 127 | 44.6% |
| Taking Aspirin | Yes | 58 | 20.4% |
|  | No | 227 | 79.6% |

**Figure 5 5.**
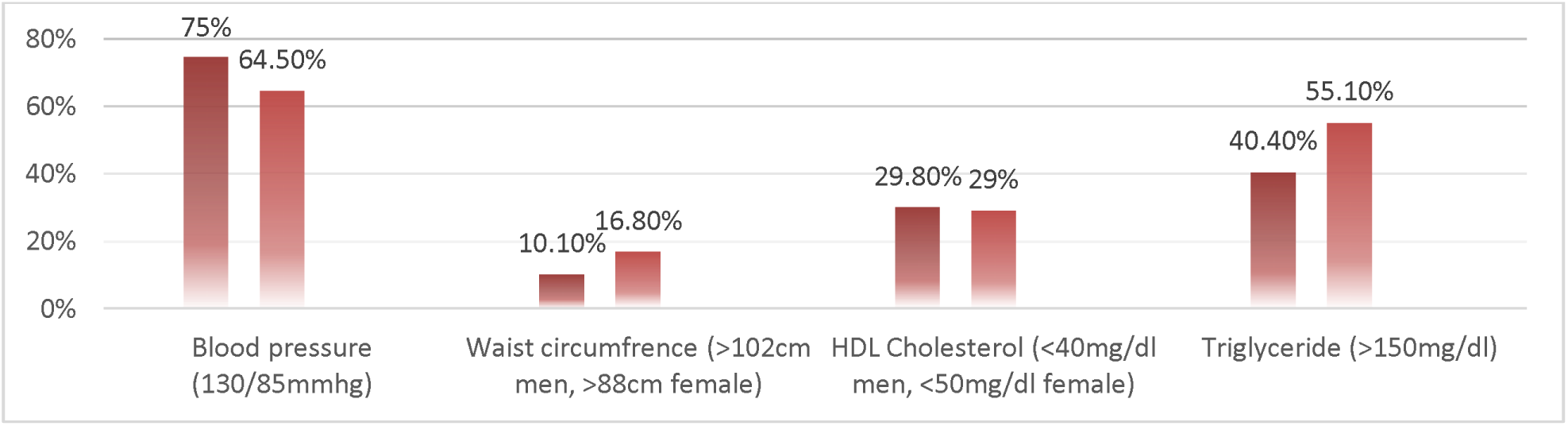
Frequency of Metabolic syndrome components among different sex among participants at ARTH, Oromia, Ethiopia from July to September 2022 EC. n(285)

### 5.8 Factors associated with Metabolic syndrome in type II DM patients

Binary logistic regression analysis was done. Smoking status was not entered into the regression analysis because of the small sample size of current smokers. Variables that transferred from the bivariate analysis into the multi-variable analysis were; Treatment for Hypertension, Lipid lowering treatment, Frequency of consuming milk and milk products occupational status, Frequency of taking Meat, Alcohol intake, Weight, Glycemic control FBS (mg/dl), Total Cholesterol (mg/dl) and LDL Cholesterol (mg/dl). After adjusting for confounders; Treatment for Hypertension, Frequency of taking Meat, Alcohol intake, Total Cholesterol (mg/dl) and LDL Cholesterol (mg/dl) had a significant association with Metabolic syndrome.

Metabolic syndrome was 2.25 times more likely to occur in participants who were taking Anti-Hypertensive medication than those who were not taking (AOR = 2.25; 95% CI = 1.177-4.279). Study participants who were Meat on daily to weekly base had 2.47 times higher odds of Metabolic syndrome as compared to those who were not taking weekly (AOR = 2.47; 95% CI = 1.323-4.6). Patients who were drinking Alcohol were 2.79 times more likely to develop Metabolic syndrome as compared to who were not (AOR = 2.79; 95% CI = 1.146-6.806). Regarding lipid profile, compared to patients who had low total cholesterol level those who had total Cholesterol more than 200mg/dl had 2.72 times more chance of developing Metabolic syndrome (AOR = 2.72; 95% CI = 1.023-7.224) and those who had LDL Cholesterol level greater than 130 were 2.31 times at risk to develop Metabolic syndrome compared to those who had low LDL Cholesterol level (AOR = 2.31; 95% CI = (1.029-5.161).

**Table 5 12.** Factors associated with metabolic syndrome among type 2 diabetes mellitus patients from Asella Referral and teaching Hospital Oromia region, Ethiopia, 2022 (n= 285)

| Variable | Category | Metabolic syndrome |  | COR (95%CI) | P-value | AOR (95%CI) | P-value |
|---|---|---|---|---|---|---|---|
|  |  | Yes | No |  |  |  |  |
| Treatment for Hypertension | Yes | 109 | 24 | 2.033 (1.161-3.559) | 0.013 | 2.245 (1.177-4.279) | **0.014** |
|  | No | 105 | 47 | 1 |  |  |  |
| Lipid lowering treatment | Yes | 123 | 35 | 1 |  |  |  |
|  | No | 91 | 36 | 0.719 (0.42-1.232) | 0.23 | 1.1087 (0.489-1.664) | 0.741 |
| Frequency of consuming milk and milk products | Daily | 54 | 15 | 1 |  |  |  |
|  | Weekly | 122 | 49 | 0.663 (0.247-1.782) | 0.415 | 0.79 (0.264-2.362 | 0.673 |
|  | Didn’t take | 38 | 7 | 0.459 (0.192-1.097) | 0.08 | 0.453 (0.162-1.265) | 0.131 |
| Frequency of taking Meat | Daily - Weekly | 134 | 34 | 1.823 (1.06-3.133) | 0.03 | 2.466 (1.323-4.6) | **0.005** |
|  | Didn’t take | 80 | 37 | 1 |  |  |  |
| Alcohol intake | Yes | 57 | 8 | 2.859 (1.290-6.335) | 0.01 | 2.793 (1.146-6.806) | **0.024** |
|  | No | 157 | 63 |  |  |  |  |
| Weight |  | 74 | 70 | 1.025 (1.001-1.049) | 0.042 | 1.018 (0.991-1.047) | 0.189 |
| Glycemic control FBS (mg/dl) | ≥130 | 124 | 35 | 1.417 (0.827-2.429) | 0.205 | 1.309 (0.714-2.399) | 0.384 |
|  | <130 | 90 | 36 | 1 |  |  |  |
| Total Cholesterol (mg/dl) | ≥200 | 57 | 6 | 3.933 (1.616-9.573) | 0.003 | 2.719 (1.023-7.224) | **0.045** |
|  | <200 | 157 | 65 | 1 |  |  |  |
| LDL Cholesterol (mg/dl) | ≥130 | 77 | 10 | 3.428 (1.661-7.076) | 0.001 | 2.305 (1.029-5.161) | **0.042** |
|  | <130 | 137 | 61 | 1 |  |  |  |

## 6 Discussion

This study was aimed at describing the magnitude of Metabolic syndrome and its associated factors among type two diabetic patients attending a Asella Referral and teaching hospital diabetic follow-up clinic. The main finding of the present study demonstrates that Metabolic syndrome is a major health concern for diabetic patients in Arsi zone and as well in Ethiopia. The associated factors like participants frequency of taking Meat, Alcohol intake, total Cholesterol level and LDL Cholesterol level are significantly associated with Metabolic syndrome.

The magnitude of Metabolic syndrome in this study was 75.1% using NCEP-ATP III criteria. The finding of this study is in line with study done in Hawassa University Comprehensive Specialized Hospital which shows the magnitude to be 70.1% using NCEP-ATP III criteria(21). Compared to other studies done in Dessie Referral Hospital and University of Gondar the result of this study showed that the magnitude of Metabolic syndrome in type two DM patients to be higher. The study done in University of Gonder showed the Prevalence of 66.7% and from Dessie Referral Hospital the prevalence was 59.3% both using the NCEP-ATP III criteria(20)(17).

From previous study done in Napal the prevalence of metabolic syndrome in diabetes patients as per NCEP-ATP III criteria was 71%(3). There are also studies showing higher prevalence compared to this study, a study done in Komfo Anokye teaching Hospital found in Ghana found the prevalence of 90.6%(29). In another way a study done in the same country but different hospital showed 58% prevalence (3). This shows the prevalence of Metabolic syndrome could vary even in the same country. This explains the difference which was found in this study and other studies done in our country acceptable.

In the study done at Hawassa University Hospital the total participants were 319, among those 67% were males, 68.2% were Urban dwellers and 36.3% of them had BMI between 25-29.9kg/m² which is comparable to our study including 285 participants, 62.5% were males, 69.5% were Urban dwellers and 43.5% had BMI between 25-30kg/m². This might have contributed for the comparable results found in these two studies. Comparing the two studies according to the significantly associated factors, in former study Uric acid level, total cholesterol level and doing regular physical exercise were strongly associated with prevalence of Metabolic syndrome whereas in our study strongly associated variables were participants frequency of taking Meat, Alcohol intake, total Cholesterol level and LDL Cholesterol level. The discrepancy in significantly associated factors can be explained by the difference in clinical characteristics of participants (21).

Comparing this study to study done in Dessie referral hospital most of the socio-demographic characteristics were comparable to our study except the proportion of female to male participants, the proportion was higher in the study done in Dessie referral hospital whereas its lover in our study, 57.1% and 37.5% for former and later studies respectively. The significant difference between Magnitude of Metabolic syndrome in these two studies might be attributed to the difference in participants who were on Anti-Hypertensive medications which is 47% in our study and 35.8% in study done in Dessie. These difference indirectly tell us more Hypertensive patients were included in our study(20).

In a study done in University of Gondar Hospital a total of 159 participants were involved among those 59.7% were females, the study showed the prevalence of Metabolic syndrome to be 66.4% based on NCEP-ATP III. Compared to our study the prevalence was lower, the difference can be explained by the higher presence of HDL Cholesterol level (<40mg/dl for males and <50mg/dl for females) in our study 70.5% vs 31.5 noting that HDL Cholesterol level is a criteria for the diagnosis of metabolic syndrome(17).

In this study Hypertension was the commonest risk factor (70.9%), followed by HDL Cholesterol (70.5%), Triglyceride (46%) and Waist circumference (12.6%) respectively. Also, the research done in Komfo Anokye Teaching Hospital, Kumasi, Ghana showed Hypertension as the commonest risk factor. In our study central obesity and high Triglyceride level were the only components of metabolic syndrome with higher prevalence in females, Hypertension was more common in males and HDL Cholesterol level was comparable. This finding is concordant with study done in Ghana where the study showed significant association between being female and having central obesity(3).

The prevalence of Metabolic syndrome in our study was 75.1% (75.3% in males and 74.8% in females) which contradicts with other studies where the prevalence was higher in females than meles (20) (30). These differences may be attributed to lower LDL Cholesterol level found in females and difference in other clinical characteristics.

In our study we found that only 158 (55.4%) of participants were taking lipid lowering medication and 58 (20.4%) of the participants was taking Aspirin, as to the recommendation of American diabetic association lipid lowering medications are recommended are recommended to be used for all type II DM patients age above 40 years and in our study 96.1% of the participants were age above 40 years. Concerning Aspirin according to ADA the recommendation is for those with CVD history or based on ASCVD risk score(7).

On the whole, there was higher prevalence of Metabolic syndrome among type II diabetic patients in this study compared to another studies done in our country (Dessie, Gondar and Hawassa) even though there are other studies that showed even higher prevalence like Napal(20)(21)(17)(30). Through logistic regression analysis, our study has shown four factors: frequency of taking Meat, Alcohol intake, total Cholesterol level and LDL Cholesterol level are significantly associated with Metabolic syndrome with adjusted odds ratio of 2.47, 2.79, 2.72 and 2.3 respectively. Knowing the commonest components and risk factors will give a guide to prevention and treatment of type II DM patients.

## 7. Limitation of the study

The main limitation of our study was the small sample size, which may have limited the power of the results. This was a cross-sectional study which took place in one particular region and may not be extended to other regions.

## 8. Conclusion

This study showed that the magnitude of Metabolic syndrome in type II DM patients was 75.1% and through logistic regression analysis, has delineated key risk factors driving morbidity. The significantly associated were, frequency of taking Meat, Alcohol intake, total Cholesterol level and LDL Cholesterol level. The most prevalent component was Hypertension followed by HDL Cholesterol, Triglyceride and Waist circumference respectively. Our study also found that suboptimal utilization of lipid lowering medications.

## 9. Recommendation

Noting the high magnitude Metabolic syndrome among type II DM patients shown by this study, I strongly recommend optimal management of type II DM patients with guideline directed lifestyle modification and pharmacologic management. I also recommend optimal utilization of lipid lowering drugs in type II DM patients.

## Data Availability

All data produced in the present work are contained in the manuscript

## Acronyms and Abbreviations

ADA: American diabetic association
AOR: Adjusted odds ratio
ARTH: Asella Referral and Teaching Hospital
AUCHS: Arsi University College of Health Science
BMI: Body mass index
Bp: Blood pressure
BSc: Bachelor of Science
CVD: Cardiovascular disease
DDS: Dietary Diversity Score
FBG: Fasting blood glucose
HDL: High density lipoprotein
IDF: International Diabetes Federation
IR: Insulin Resistance
LDL: Low density lipoprotein
Mets: Metabolic syndrome
NCD: Noncommunicable diseases
NCEP-ATP III: National Cholesterol Education Program Adult Treatment Panel III
T II DM: Type 2 Diabetes Mellitus
TG: Triglyceride
WHO: World Health Organization

